# Mental Health Stigma and Help-Seeking among South Asian American Young Adults in Texas: A Mixed-Methods Community-Based Needs Assessment

**DOI:** 10.64898/2026.09.05.26362356

**Authors:** Sophia Khan, Xuewei Chen

## Abstract

**Background:** Although barriers to mental health help-seeking among Asian American populations are well documented, less is known about how favorable individual attitudes toward mental health treatment coexist with family and community influences among South Asian American young adults or which intervention strategies members of this population themselves prioritize.

**Methods:** A cross-sectional mixed-methods survey was completed by 191 South Asian American young adults aged 18-30 years residing in Texas between May and June 2026 using convenience sampling. Participants completed 5-point Likert-scale measures assessing mental health attitudes, stigma, help-seeking, and barriers to care. 144 participants also provided responses to optional open-ended questions exploring barriers to mental health care and recommendations for improving access to mental health support. Quantitative data were analyzed using IBM SPSS Statistics, and qualitative responses underwent thematic analysis in MAXQDA.

**Results:** Participants reported generally positive attitudes toward mental health care and high perceived importance of mental health despite family- and community-level barriers. Greater family pressure was strongly associated with increased mental health stigma (*r* = 0.711, *p* < 0.001) and perceived barriers to care (*r* = 0.654, *p* < 0.001), while greater perceived barriers were associated with less favorable help-seeking attitudes (*r* = −0.158, *p* = 0.036). Qualitative responses contextualized these findings, identifying anticipated community judgment, family expectations, privacy concerns, and limited mental health literacy among older generations as barriers to care. Participants emphasized culturally responsive care and recommended mental health education involving parents, elders, faith leaders, and trusted community institutions.

**Conclusions:** South Asian American young adults may hold favorable attitudes toward professional mental health care while navigating family, community, and cultural contexts that constrain help-seeking. Participant-generated recommendations prioritized culturally responsive care and community-level mental health education involving parents, elders, faith leaders, and trusted institutions, suggesting that interventions targeting individual attitudes alone may be insufficient.

## Introduction

Increased awareness of mental illness has improved public mental health literacy, reduced stigma, and increased willingness to seek care.^1^ However, the utilization of mental health resources remains disproportionately low among South Asian Americans.^2^ Although previous studies have identified cultural stigma and family expectations as barriers to care, less is known about how these factors influence help-seeking or what community-informed strategies may improve access to mental health services.^3^

South Asian families emphasize family honor, reputation, collectivist values, and social image, which may contribute to mental health stigma and discourage professional help-seeking.^4,5^ Despite the rapid growth of the South Asian American population, this group remains understudied and is frequently aggregated into broader Asian American populations, limiting recognition of culturally specific barriers and the development of culturally responsive interventions.^6–8^ Cultural stigma may also discourage open discussion and research participation, further limiting understanding of mental health needs within this population.^9^ Improving understanding of these culturally specific barriers is essential for developing culturally responsive interventions and improving equitable access to mental health care among South Asian Americans.

Mental health help-seeking barriers among Asian American populations have been well documented. Kim and Lee’s systematic review identified multiple individual, cultural, and structural factors associated with mental health help-seeking across diverse Asian American populations.^10^ However, findings derived from aggregated or predominantly East and Southeast Asian samples may not fully characterize South Asian-specific family, cultural, and community contexts.^7,10^ More recent South Asian-specific literature has similarly identified stigma, family expectations, privacy concerns, and culturally responsive care as important influences on mental health services access.^3,8^ Thus, an important remaining question is not simply whether barriers exist, but how favorable individual attitudes toward mental health treatment coexist with family- and community-level influences among South Asian American young adults and which strategies members of this population themselves recommend for improving access.

Young adulthood may be particularly important for examining this question because emerging autonomy and evolving attitudes toward mental health occur while family and social relationships continue to influence stigma, disclosure, and help-seeking decisions.^11^ Understanding this tension may help distinguish individual reluctance to seek treatment from social and cultural contexts that constrain help-seeking despite favorable attitudes toward professional mental health care.

Therefore, a mixed-methods study examined how family expectations, community stigma, structural barriers, and cultural values influence mental health attitudes and help-seeking among South Asian American young adults (ages 18-30 years) residing in Texas. Quantitative analyses examined relationships among these constructs, while qualitative responses explored participants’ experiences and recommendations for improving access. By integrating these approaches, this study sought to extend existing barrier-focused literature by examining the coexistence of favorable individual treatment attitudes with persistent family and community influences, while also exploring participant-generated priorities for culturally responsive interventions. The theoretical framework guiding this study is presented in Figure 1.

**Figure 1.**
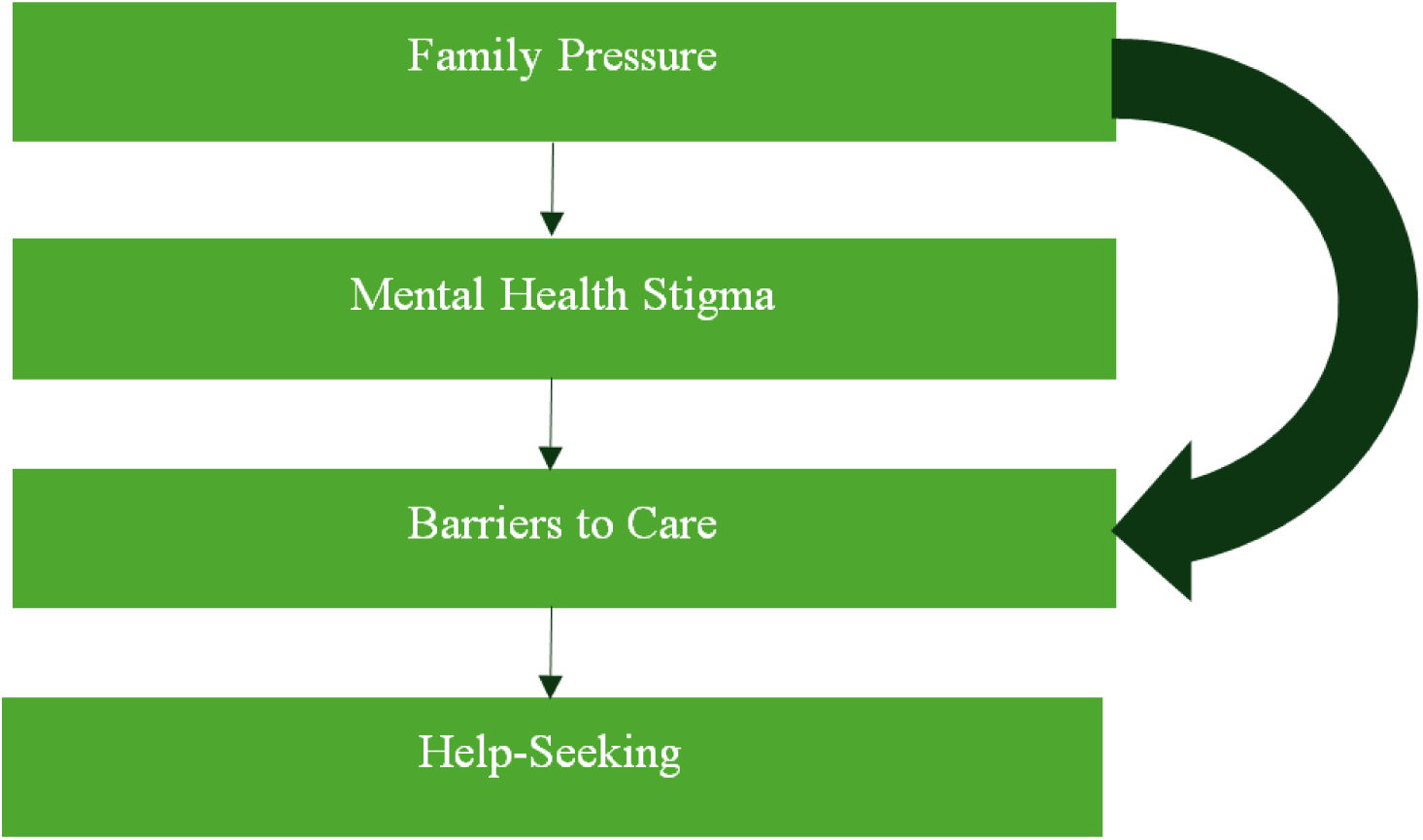
Conceptual framework illustrating hypothesized relationships among family pressure, mental health stigma, perceived barriers to care, and help-seeking among South Asian American young adults. Straight arrows represent the primary hypothesized pathway, whereas curved arrows represent additional direct associations identified in the quantitative analyses.

## Methods

### Study Design

This cross-sectional mixed-methods study used a survey consisting of quantitative Likert-scale items and optional open-ended questions to examine mental health attitudes, stigma, family and community influences, barriers to care, and participant-recommended strategies to improve culturally responsive care among South Asian American young adults (ages 18-30) residing in Texas. A convergent mixed-methods design was used to integrate quantitative survey findings with qualitative responses. The survey instrument was reviewed by the research team for clarity and content validity before distribution.

### Participants

Eligible participants were South Asian American young adults aged 18-30 years residing in Texas. No formal a priori sample size calculation was performed. Participants were recruited during the predefined study period from May to June 2026. Of 227 survey responses received, responses from individuals who did not provide consent, did not meet eligibility criteria, or had incomplete survey data were excluded, resulting in a final analytic sample of 191 participants. Among these participants, 144 completed the optional open-ended survey questions and were included in the qualitative analysis.

### Recruitment

Participants were recruited using convenience sampling from South Asian community organizations, religious organizations, university student organizations, community events, and social media between May and June 2026.

### Study Setting

This study was conducted among South Asian American communities in Texas between May and June 2026. Recruitment occurred through South Asian community organizations, religious organizations, university student organizations, community events, and social media. Participants resided throughout Texas, with recruitment primarily representing major metropolitan areas including Dallas-Fort Worth, Houston, and Austin.

### Data Collection

Data were collected using paper-based surveys distributed at community events and electronic surveys administered through REDCap^12,13^ between May and June 2026. Participation was anonymous and voluntary. Participants reviewed and provided informed consent electronically or in writing before survey participation.

### Measures

The surveys consisted of demographic questions and multiple 5-point Likert-scale items assessing mental health attitudes, family and cultural influences, mental health stigma, help-seeking behaviors, and perceived barriers to care (Table 1). Responses for the mental health attitudes, family and cultural influence, stigma, and help-seeking domains ranged from strongly disagree to strongly agree, whereas responses for perceived barriers ranged from not a barrier to a major barrier. Because no single validated instrument comprehensively assessed the study objectives, the survey instrument was developed specifically for the present study following a review of literature on Asian American mental health stigma, help-seeking, family and community influences, and barriers to culturally responsive mental health care.^14–17^ The complete English-language survey instrument is provided in Additional file 1. Two optional open-ended questions explored participants’ experiences with mental health, barriers, and recommendations for improving access to culturally responsive care.

**Table 1.** Survey Domains, Constructs Measured, Example Items, Response Scales, and Number of Items.

|  |  |  |  |  |
| --- | --- | --- | --- | --- |
| Survey Domain | Construct Measured | Example Question(s) | Response Scale | Number of Items |
| Eligibility | Study eligibility | "Are you between the ages of 18-30?" "Do you identify as South Asian?" "Do you reside in Texas?" | Yes/No | 3 |
| Demographics | Participant characteristics | Age, gender, ethnicity, education/employment, city of residence | Multiple choice | 5 |
| Mental Health Attitudes | General perceptions of mental health | "Mental health is an important part of overall health"; "Therapy is a valid treatment" | 5-point Likert (1=Strongly agree – 5=Strongly disagree) | 4 |
| Family and Cultural Influence | Family support and community expectations | "My family would support me if I sought treatment"; "There is pressure in my family to maintain a positive image" | 5-point Likert | 4 |
| Mental Health Stigma | Community stigma and internalized beliefs | “Seeking mental health care is seen as a sign of weakness”; “Mental health struggles are often kept private within families | 5-point Likert | 4 |
| Help-Seeking Behavior | Attitudes toward seeking professional care | “I would feel comfortable seeking professional mental health care”; “I trust mental health professionals”; “I would prefer a provider who understands my cultural background | 5-point Likert | 4 |
| Barriers to Care | Perceived barriers to accessing care | Cost, culturally competent providers, language, family expectations, fear of judgment, awareness of resources, religious/cultural beliefs | 5-point barrier scale (1=Not a barrier – 5=Major barrier) | 7 |
| Open-ended Questions | Lived experiences and recommendations | “What are the biggest barriers?” “What changes would make it easier to seek support?” | Free text | 2 |

Composite scores were calculated by averaging conceptually related items including family pressure, mental health stigma, perceived barriers to care, and help-seeking attitudes (Table 2). Composite scales were developed conceptually rather than according to the survey section. The primary mental health stigma composite consisted of six items drawn from the mental health stigma and family/cultural influence domains. Because two items overlapped with the family pressure composite, a four-item non-overlapping stigma composite was used in the multivariable regression model in which family pressure and stigma were entered simultaneously. The non-overlapping stigma composite demonstrated acceptable internal consistency (Cronbach’s α = 0.708). Positively worded items assessing mental health importance, therapy importance, family openness and support, comfort seeking care, knowledge of services, trust in professionals, and preference for culturally informed providers were reverse coded before composite score calculation so that higher scores consistently reflected greater family pressure, mental health stigma, perceived barriers, and more positive help-seeking attitudes.

**Table 2.** Reliability and Descriptive Statistics for Composite Scales.

| **Table 2. Reliability and Descriptive Statistics for Composite Scales.** |  |  |  |  |
| --- | --- | --- | --- | --- |
| Scale | Number of Items | Cronbach’s $\alpha$ coefficient | Mean | SD |
| Family pressure | 4 | 0.728 | 3.12 | 0.87 |
| Mental Health Stigma | 6 | 0.743 | 3.79 | 0.69 |
| Barriers to Care | 6 | 0.836 | 2.37 | 0.93 |
| Help-Seeking Score | 2 | 0.778 | 4.07 | 0.82 |
| Note: A four-item non-overlapping mental health stigma composite (Cronbach’s $\alpha = 0.708$) was used in the multivariable regression to avoid item overlap with the family pressure composite. | | | | |

### Statistical Analysis

Quantitative analyses were performed using IBM SPSS Statistics for Mac, Version 28.0 (IBM Corp., Armonk, NY, USA). Responses that did not meet the study’s completeness criteria were excluded prior to analysis; remaining item-level missing values were not imputed. Descriptive statistics summarized participant demographics and survey responses, and internal consistency of composite scales was assessed using Cronbach’s alpha. Pearson correlation analyses examined associations among the composite measures of family pressure, mental health stigma, perceived barriers to care, and help-seeking attitudes, including the relationships between family pressure and mental health stigma, family pressure and perceived barriers to care, mental health stigma and perceived barriers to care, and help-seeking attitudes with each of the other composite measures. Chi-square tests examined associations between demographic variables (age group, gender, ethnicity) and selected survey responses, including perceived pressure to maintain family image, comfort seeking professional mental health care, and beliefs that mental health concerns are taken seriously by others. One-way analysis of variance (ANOVA) with post hoc testing compared help-seeking attitude across age groups. Multiple linear regression evaluated the adjusted associations of family pressure, mental health stigma, perceived barriers to care, and age group with help-seeking attitudes. To avoid item overlap between family pressure and mental health stigma in the multivariable model, the four-item non-overlapping stigma composite was used for this analysis. Multicollinearity was assessed using variance inflation factors (VIFs) and tolerance statistics.

### Qualitative Analysis

Responses were coded inductively in MAXQDA Analytics Pro. Codes were iteratively refined and organized into broader themes that captured recurring patterns across participant responses, following Braun and Clarke’s six-phase approach.^18^

### Potential Sources of Bias

To reduce potential social desirability bias surrounding sensitive mental health topics, survey participation was anonymous. Participants were recruited through multiple community, religious, university, and online settings to broaden recruitment. However, convenience sampling may have introduced selection bias, and these limitations are considered in the Discussion.

### Ethics

This study was approved by the Sam Houston State University Institutional Review Board (IRB #2026278). All participants provided informed consent. Survey responses were anonymous and contained no personal identifiable information. Contact information provided for optional focus groups was stored separately from survey data. This study was conducted in accordance with the Declaration of Helsinki.

## Results

### Quantitative Findings

A total of 191 South Asian American young adults in Texas completed the survey. Most participants were 21-23 years old (60.7%), female (68.6%), and identified as Indian (58.1%) or Pakistani (35.1%). Many participants were graduate students (40.8%), and nearly half resided in the Dallas-Fort Worth metropolitan area (49.2%). Participant characteristics are presented in Table 3.

**Table 3.** Demographic Characteristics of Study Participants (*N* = 191).

| **Table 3. Demographic Characteristics of Study Participants (N = 191).** |  |
| --- | --- |
| **Characteristic** | **N (%)** |
| **Age** |  |
| 18-20 years | 26 (13.6%) |
| 21-23 years | 116 (60.7%) |
| 24-26 years | 43 (22.5%) |
| 27-30 years | 4 (2.1%) |
| missing | 2 (1.0%) |
| **Gender** |  |
| Female | 131 (68.6%) |
| Male | 58 (30.4%) |
| Nonbinary | 1 (0.5%) |
| missing | 1 (0.5%) |
| **Ethnicity** |  |
| Indian | 111 (58.1%) |
| Pakistani | 67 (35.1%) |
| Other South Asian | 10 (5.2%) |
| missing | 3 (1.6%) |
| **Education/Employment** |  |
| Undergraduate Student | 34 (17.8%) |
| Graduate Student | 78 (40.8%) |
| Employed Full-Time | 51 (26.7%) |
| Employed Part-Time | 11 (5.8%) |
| Other | 12 (6.3%) |
| Prefer not to answer | 4 (2.1%) |
| missing | 1 (0.5%) |
| **City of residence** |  |
| Dallas-Fort Worth | 94 (49.2%) |
| Houston | 38 (19.9%) |
| Austin | 30 (15.7%) |
| San Antonio | 6 (3.1%) |
| Other Texas | 6 (3.1%) |
| missing | 17 (8.9%) |

Participants reported favorable attitudes toward professional mental health care. For agreement items, lower mean scores indicate greater agreement because responses were coded from 1 = Strongly agree to 5 = Strongly disagree. Mental health was viewed as an important component of overall health (M = 1.14, SD = 0.36), and therapy and counseling were considered a valid approach to addressing mental health concerns (M = 1.51, SD = 0.64). Participants generally reported trusting mental health professionals (M = 1.97, SD = 0.89). The composite Help-Seeking Attitudes score indicated positive attitudes toward professional mental health care (M = 4.07, SD = 0.82). However, participants also reported that mental health struggles were often kept private within families (M = 1.64, SD = 0.70), highlighting the persistence of stigma despite generally positive attitudes toward treatment. Individual survey items are shown in Table 4. Mean ratings for perceived barriers to mental health help-seeking are shown in Figure 2.

**Figure 2.**
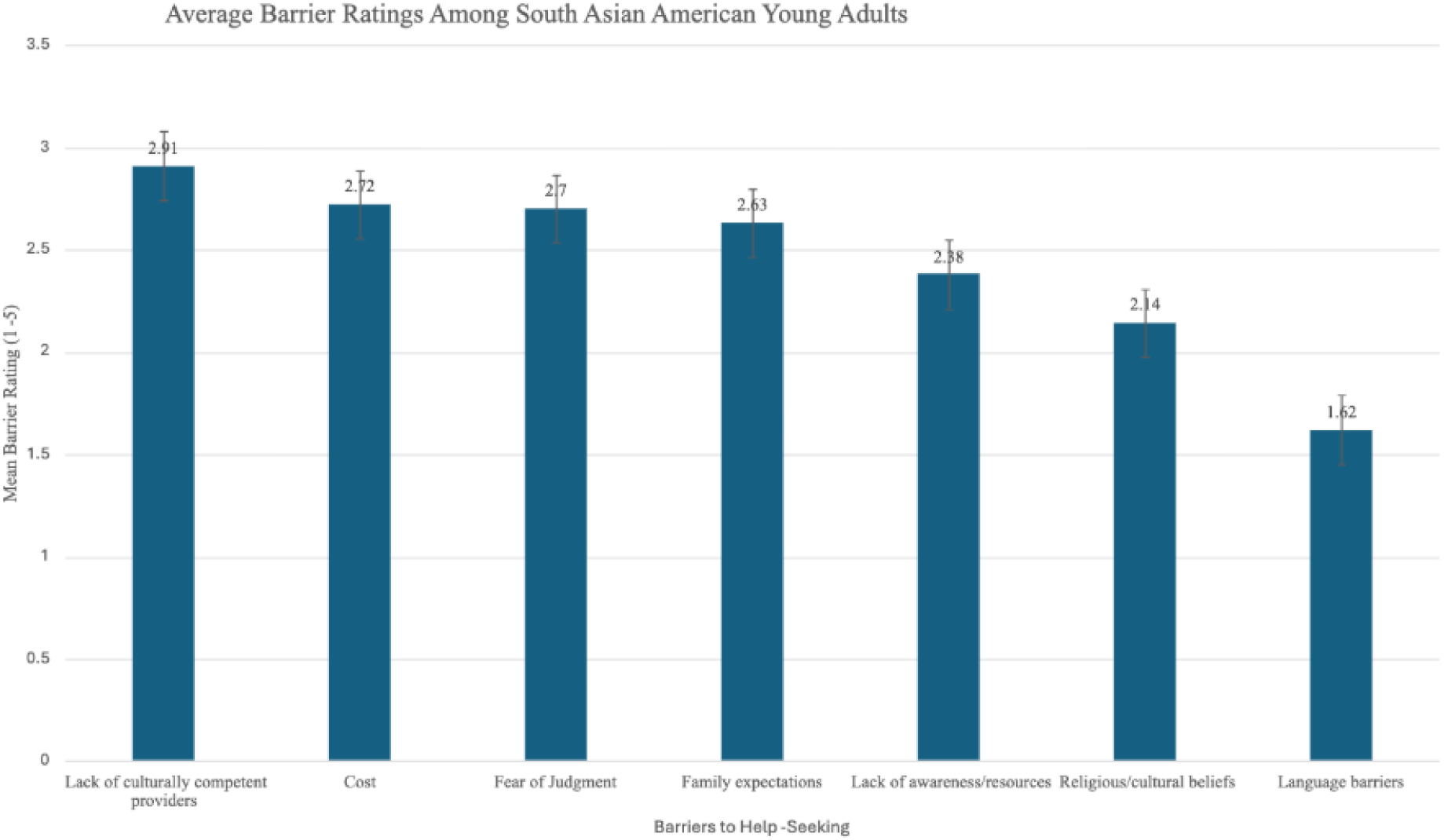
Mean ratings of perceived barriers to mental health help-seeking among South Asian American young adults.

**Table 4.** Descriptive Statistics for Individual Survey Items by Study Domain.

|  |  |  |
| --- | --- | --- |
| **Survey Item** | **Mean** | **SD** |
| **Mental Health Attitudes** |  |  |
| Mental health is an important part of overall health | 1.14 | 0.36 |
| I feel comfortable talking about mental health with others | 2.08 | 0.92 |
| Mental health issues are taken seriously by people around me | 2.38 | 0.95 |
| Therapy or counseling is a valid way to address mental health concerns | 1.51 | 0.64 |
| **Family and Community Influences** |  |  |
| In my family, mental health is openly discussed | 2.94 | 1.24 |
| My family would support me if I sought mental health treatment | 2.20 | 1.09 |
| There is pressure in my family to maintain a positive image | 2.02 | 0.97 |
| I worry about what others in my community would think if I sought help | 2.84 | 1.17 |
| **Mental Health Stigma** |  |  |
| There is stigma around mental health in my community | 1.85 | 0.80 |
| Seeking mental health care is seen as a sign of weakness | 2.88 | 1.32 |
| People in my community are hesitant to talk about mental health | 1.97 | 0.87 |
| Mental health struggles are often kept private within families | 1.64 | 0.70 |
| **Help-Seeking** |  |  |
| I would feel comfortable seeking professional mental health care | 1.88 | 0.92 |
| I know how to access mental health services | 1.94 | 0.99 |
| I trust mental health professionals | 1.97 | 0.89 |
| I would prefer a provider who understands my cultural background | 1.57 | 0.75 |
| **Barriers to Care** |  |  |
| Cost/financial concerns | 2.72 | 1.25 |
| Lack of culturally competent providers | 2.91 | 1.14 |
| Language barriers | 1.62 | 1.12 |
| Family expectations | 2.63 | 1.32 |
| Fear of judgment | 2.70 | 1.25 |
| Lack of awareness or resources | 2.38 | 1.26 |
| Religious or cultural beliefs | 2.14 | 1.32 |

**Note.** Agreement items were rated on a 5-point Likert scale (1 = Strongly agree; 5 = Strongly disagree). Barrier items were rated from 1 = Not a barrier to 5 = Major barrier.

Pearson correlation analyses showed that family pressure was strongly correlated with mental health stigma (*r* = 0.711, *p* < 0.001) and perceived barriers to mental health care (*r* = 0.654, *p* < 0.001). Stigma was modestly associated with barriers to care (*r* = 0.273, *p* < 0.001), while perceived barriers were negatively associated with help-seeking attitudes (*r* = −0.158, *p* = 0.036). The full correlation matrix is shown in Table 5.

**Table 5.**
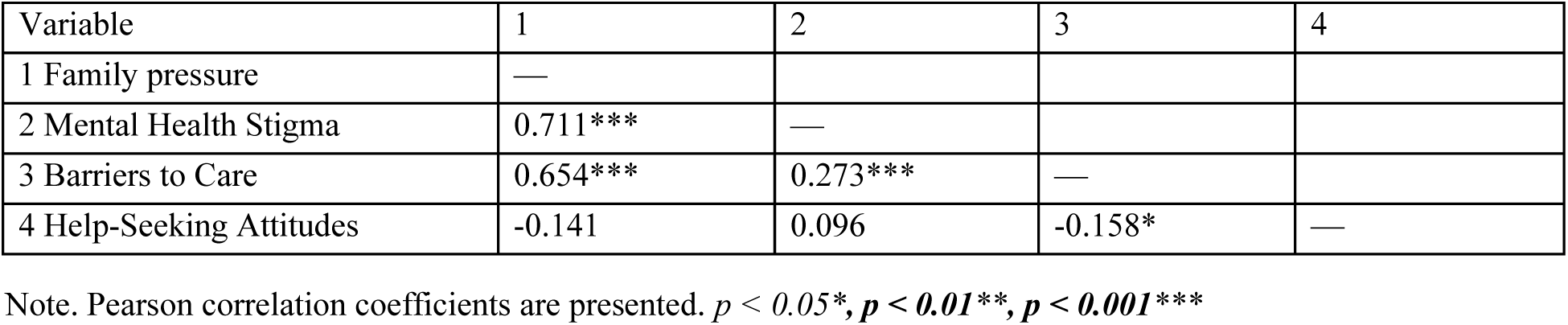
Pearson Correlation Matrix of Composite Study Variables.

| **Table 5. Pearson Correlation Matrix of Composite Study Variables.** |  |  |  |  |
| --- | --- | --- | --- | --- |
| Variable | 1 | 2 | 3 | 4 |
| 1 Family pressure | — |  |  |  |
| 2 Mental Health Stigma | 0.711*** | — |  |  |
| 3 Barriers to Care | 0.654*** | 0.273*** | — |  |
| 4 Help-Seeking Attitudes | -0.141 | 0.096 | -0.158* | — |

Note. Pearson correlation coefficients are presented. $p < 0.05^*$ , $p < 0.01^{**}$ , $p < 0.001^{***}$

Help-seeking attitudes differed significantly across age groups, with participants aged 24-26 reporting significantly higher help-seeking attitudes than participants aged 18-20 (*p* = 0.007, *η*² = 0.055). Female participants were significantly more likely than male participants to report comfort seeking professional mental health care (*χ*²(8) = 18.01, *p* = 0.021). Additionally, perceptions that mental health is taken seriously differed across age groups (*χ*² (12) = 27.67, *p* = 0.006), with older participants reporting that mental health was taken more seriously by those around them than younger participants. Family pressure did not differ significantly across age groups or between Indian and Pakistani participants. Compared to Indian participants, Pakistani participants reported greater concerns regarding community stigma, privacy, hesitation discussing mental health, and community perceptions of help-seeking (Table 6).

**Table 6.** Group Comparisons Across Demographic Characteristics.

| Table 6. Group Comparisons Across Demographic Characteristics |  |  |  |
| --- | --- | --- | --- |
| Variable | Comparison | Statistic | $p$ |
| Help-Seeking Score | Age group | $F(3, 184) = 3.59$ | 0.015 |
| Family Pressure Score | Indian vs. Pakistani | $t(176) = 1.41$ | 0.159 |
| Pressure to Maintain Family Image | Ethnicity | $\chi^2(20) = 74.65$ | <0.001 |
| Help-Seeking Comfort | Gender | $\chi^2(8) = 18.01$ | 0.021 |
| Belief Mental Health is Taken Seriously | Age Group | $\chi^2(12) = 27.67$ | 0.006 |

Note. $\eta^2$ = eta squared

In the multivariable model using the non-overlapping mental health stigma composite, mental health stigma and age group were significantly associated with help-seeking after adjustment for the other included predictors. Family pressure and perceived barriers to care were not independently significant. The overall model was statistically significant, F (4, 182) = 6.40, p < 0.001, and explained 12.3% of the variance in help-seeking attitudes (R^2^ = 0.123; adjusted R^2^ = 0.104)

| Predictor | B | SE | $\beta$ | 95% CI for B | $p$ |
| --- | --- | --- | --- | --- | --- |
| Family pressure | -0.169 | 0.098 | -0.182 | -0.361 to 0.024 | 0.086 |
| Mental Health Stigma | 0.255 | 0.095 | 0.219 | 0.067 to 0.443 | 0.008 |
| Barriers to Care | -0.120 | 0.082 | -0.138 | -0.281 to 0.041 | 0.144 |
| Age Group | 0.244 | 0.087 | 0.199 | 0.072 to 0.417 | 0.006 |

Note. B = unstandardized regression coefficient; SE = standard error; $\beta$ = standardized regression coefficient; CI = confidence interval. Model statistics: $R^2 = 0.123$ , Adjusted $R^2 = 0.104$ , $F(4, 182) = 6.40$ , $p < 0.001$ . VIF values were below 2.30.

To better understand the cultural and interpersonal experiences underlying these quantitative relationships, qualitative responses were analyzed using inductive thematic analysis. Seven interconnected themes emerged that further explained participants’ individual experiences with mental health attitudes and help-seeking experiences.

### Qualitative Findings

Seven themes emerged from the qualitative analysis: (1) mental health stigma remains a central barrier to care; (2) family functions as both a barrier and facilitator to mental health care; (3) intergenerational differences shape mental health attitudes; (4) mental health literacy influences recognition of emotional distress and help-seeking; (5) community judgment, family image, and privacy concerns influence help-seeking; (6) the need for culturally responsive, patient-centered care; and (7) community-based solutions are viewed as essential for improving mental health access.

#### Theme 1. Mental health stigma remains a central barrier to care

Participants repeatedly described mental health stigma as one of the most significant barriers to seeking mental health care. Stigma was viewed as deeply embedded within South Asian family and community culture, shaping attitudes toward mental health and discouraging open discussion. Many participants reported that emotional concerns were minimized, dismissed, or avoided altogether, creating feelings of shame and hesitation to disclose struggles or seek professional support.

Representative quote: “However, there is still a lot of resistance to bringing up these problems (“log kya kahenge”, what will people say) or when they do come up, talking around the issue instead of talking about it openly. The community needs to be more open and understanding of mental health issues, they need to be more normalized.”

#### Theme 2. Family functions as both a barrier and facilitator to mental health care

Participants consistently described family, particularly parents, as a major influence in shaping mental health attitudes and help-seeking. Many reported that parents minimized emotional concerns, viewed mental health differently from younger generations, or wished to remain involved in treatment decisions, contributing to fear of parental judgment and reluctance to seek professional support. However, participants also described families as potential facilitators of care, with greater parental openness and understanding associated with increased comfort discussing mental health and seeking treatment. These findings suggest that families play a central role in reinforcing or reshaping mental health beliefs.

Representative quotes: “What really helps me is that my parents value mental health now. I got diagnosed with BPD years ago and since then they’ve gotten better and better at understanding that I need extra support. They push me to get professional help, I am able to talk openly with them about it and they even take classes about BPD. They understand mental health is a huge part of their child’s wellbeing and safety.”

“Families try to erase mental health struggles to avoid admitting their child may have disabilities, both due to personal grief and to maintain reputation. Mental health issues are seen as personal failings that can be remedied through motivation, positivity, praying, or marriage and having kids. Therapists who may encourage boundaries or moving out of home are seen as threats.”

#### Theme 3. Intergenerational differences shape mental health attitudes

Participants consistently described differences in mental health attitudes between younger and older generations. Younger generations were viewed as more accepting of mental health as an important component of overall health and more supportive of help-seeking. In contrast, older generations were described as minimizing emotional distress and interpreting psychological struggles through personal responsibility or life experience rather than mental illness. As a result, participants reported that emotional concerns were frequently dismissed or overlooked within families, contributing to persistent stigma and influencing help-seeking attitudes.

Representative quote: “Older generations are often unable to understand that mental health concerns are not comparative. Just because they have experienced significant hardship does not mean younger generations’ mental health concerns are not valid.”

#### Theme 4. Mental health literacy influences recognition of emotional distress and help-seeking

Participants consistently described mental health literacy as an important influence on recognizing emotional distress and deciding when to seek professional support. Rather than lacking awareness of available mental health services, many participants described difficulty recognizing when emotional distress warranted professional intervention. Emotional concerns were often minimized, normalized, or dismissed until they became severe, delaying treatment. Participants emphasized that improving mental health literacy within families and communities could increase recognition of mental health concerns, reduce stigma, and encourage earlier help-seeking. Many also compared mental and physical health, noting that emotional concerns were often given less recognition or urgency despite viewing both as equally important.

Representative quote: “I first started showing signs of depression, I was told to pray more and meditate, and it would simply go away. But that is how older generations were taught about how to deal with mental health. It is not a transient feeling; your mental health is always with you, and it is a major component of your health and can even manifest as other issues”

#### Theme 5. Community judgment, family image, and privacy concerns influence help-seeking

Participants described community judgment, concerns regarding family reputation, and privacy as important barriers to seeking mental health care. Many explained that mental health struggles were viewed through a lens of shame or weakness, contributing to fears of negative judgment and discouraging open discussion or professional help-seeking. Because many South Asian communities are closely connected, participants also expressed concerns about confidentiality, gossip, and others becoming aware of their mental health treatment. Participants emphasized that maintaining privacy was not simply a matter of convenience but a way to protect themselves and their families from anticipated community judgment.

Representative quotes: “It’s the idea that if you have mental health issues, it looks bad not only on yourself but on your family as well, and it becomes a topic of gossip. I believe this worsens mental health, especially among South Asian youth and young adults, and should be addressed to improve standards of care.”

#### Theme 6. Need for culturally responsive, patient-centered care

Participants consistently emphasized the importance of culturally responsive, patient-centered mental health care. Many described wanting providers who understood South Asian cultural values, family dynamics, religious influences, and community-specific barriers to help-seeking. Although several participants believed that increasing the number of South Asian mental health providers could improve representation and trust, many emphasized that cultural concordance was not essential. Instead, participants valued providers who demonstrated cultural humility, openness, and a willingness to understand South Asian experiences, suggesting that culturally informed care was more important than provider ethnicity alone.

Representative quotes: “To build trust in medical professionals in the mental health field, more professionals who share a cultural or ethnic background with the target populations (i.e., South Asian professionals) and/or training in cultural competency may help.”

#### Theme 7. Community-based solutions are viewed as essential

Participants consistently emphasized that improving mental health access would require interventions extending beyond individual help-seeking and healthcare settings. Many advocated for community-based education to improve mental health literacy, reduce stigma, and increase treatment acceptance among parents and older generations.

Religious institutions, including churches, mosques, temples, and other places of worship, were frequently identified as trusted settings for delivering mental health education. Participants also emphasized the influential role of parents, elders, and respected community leaders in shaping cultural attitudes, suggesting that community engagement and intergenerational education are necessary for improving mental health access within South Asian communities.

Representative quotes: “I think better screening and destigmatizing mental health issues could make it easier. I also think creating spaces for older generations to understand that the things they have experienced may have affected them would make it easier for them to understand how younger generations may be struggling. Ultimately, I think the root of a lot of mental health issues in South Asian communities comes from the invalidation of lived experiences, and I think creating spaces where these experiences are validated can help individuals realize that they have struggled and it’s okay for them to have struggled.”

### Integration of Quantitative and Qualitative Findings

The qualitative findings provided important context for the quantitative relationships observed in this study. Although participants generally reported positive attitudes toward professional mental health care and recognized mental health as an important component of overall health, many described persistent family and community stigma surrounding mental health concerns. These findings are consistent with the quantitative results demonstrating that greater family pressure was associated with increased mental health stigma and perceived barriers to care, suggesting that positive individual attitudes alone may be insufficient to overcome family and cultural influences on help-seeking.

Qualitative findings provided additional context for the relationships among family pressure, stigma, and help-seeking. Although family pressure was not independently associated with help-seeking attitudes in the multivariable model, participants described concerns surrounding community judgment, privacy, differing perspectives between generations, family reputations as important influences on decisions to seek professional support. Participants emphasized that improving mental health access would require culturally responsive care, greater mental health literacy, and community-based education involving parents, elders, and trusted community leaders. Overall, the quantitative and qualitative findings suggest that improving mental health access among South Asian American young adults requires interventions that extend beyond individual attitudes to address broader family, cultural, and community influences.

## Discussion

This mixed-methods study found that, although participants reported positive attitudes toward therapy and the importance of mental health, family expectations, community stigma, privacy concerns, and cultural beliefs remained important influences on help-seeking among South Asian American young adults in Texas. Interestingly, mental health stigma was not significantly associated with help-seeking attitudes in the bivariate analysis but demonstrated a positive association in the multivariable model using the non-overlapping stigma composite.

Because this association emerged only after adjustment for family pressure, perceived barriers, and age, it may reflect shared variance or suppression among these related psychosocial constructs rather than a direct positive relationship between stigma and help-seeking. This adjusted association should therefore be interpreted cautiously and does not indicate that greater stigma promotes help-seeking. Qualitative findings further illustrated how family and community contexts shaped decisions surrounding disclosure and treatment while identifying culturally responsive care and community-based interventions as key strategies for improving mental health access. Overall, these findings suggest that improving mental health access among South Asian American young adults requires interventions that address not only individual attitudes but also the broader family, cultural, and community contexts in which help-seeking decisions occur.

Consistent with previous research, Asian Americans generally report positive attitudes toward mental health counseling despite disproportionately low utilization of mental health services.^19, 20^ Among South Asian communities, concerns regarding family reputation and expectations have been identified as important barriers to help-seeking.^21^ Building on previous research, this mixed-methods study examined family- and community-level influences on help-seeking among South Asian American young adults in Texas while integrating participant-informed recommendations for improving culturally responsive mental health care.^3,22,23^ The focus on young adults (18-30 years) may also help explain why participants reported positive attitudes toward mental health despite persistent family- and community-level barriers, reflecting a developmental period characterized by increasing independence and evolving perspectives on mental health.

One of the principal findings of this study was that family serves as both a barrier and a facilitator for mental health care. While family pressure was not independently associated with help-seeking in the revised multivariable model, its strong bivariate associations with stigma and perceived barriers, together with the qualitative findings, suggest that family influences remain an important component of the broader help-seeking context. Qualitative responses illustrate how family interactions shaped disclosure, treatment acceptance, and help seeking. Although many participants described invalidation of emotional concerns and pressure to maintain family image, others reported increasing family support following greater mental health awareness, personal experiences, and open discussion. Together, these findings suggest that family influence is dynamic and may change with greater mental health literacy and openness to discussion.

These findings are consistent with previous research demonstrating that family can either support or hinder mental health adjustment and service utilization among South, East, and Southeast Asian populations.^24^ Other studies have similarly identified family conflict, limited mental health literacy, and family-based interventions as important influences on access to care.^25^ Nationally representative data have also shown that family conflict is associated with greater use of formal and informal mental health services, whereas family cohesion is associated with increased family support among first-generation Asian Americans.^26^ Unlike much of existing literature, participants from the study described family attitudes as evolving over time rather than remaining fixed, suggesting that family- and community-centered mental health education may represent a promising strategy for improving access to care.

Participants consistently emphasized that culturally responsive care was essential for building trust in mental health services. Although many participants preferred South Asian mental health professionals, they also valued providers who demonstrated cultural humility and a willingness to understand community-specific experiences. These findings suggest that culturally responsive communication and provider training may improve trust and treatment engagement even when provider-patient ethnic concordance is not possible. Clinicians should recognize that reluctance to seek care may reflect concerns regarding privacy, family expectations, and anticipated community judgment rather than opposition to mental health treatment. Exploring family context, discussing confidentiality early, and using culturally responsive communication may therefore improve engagement.

Participants’ emphasis on cultural humility over ethnic matching aligns with growing evidence suggesting that therapist cultural competence may be more important than shared racial or ethnic identity alone. A 2026 mixed-methods study of 86 ethnically diverse college students found that although approximately half preferred a therapist of the same race or ethnicity, cultural competence and humility were prioritized.^27^ The APA defines cultural humility as an ongoing process of self-reflection, openness, and awareness of power dynamics that fosters respectful therapeutic relationships.^28^ Similarly, a population-based study found that although Asian Americans preferred providers who shared or understood their culture, relatively few were able to access such providers, highlighting the importance of clinicians demonstrating cultural humility.^29^ Overall, participants themselves distinguished cultural humility from ethnic concordance, suggesting that openness and willingness to understand South Asian family dynamics may be more important than shared ethnicity alone for establishing trust in mental health care.

The present findings regarding privacy and anticipated community judgment are consistent with previous literature. Indian immigrants have described reluctance to seek care because of confidentiality concerns within close-knit communities.^30^ A systematic review found that treatment engagement improved when clinicians explored patients’ cultural perspectives and tailored communication to their preferences rather than relying on standardized approaches.^31^ This recommendation is consistent with research on broaching, the DSM-5-TR Cultural Formulation Interview (CFI), and the APA Resource Document on Race and Racism, all of which encourage clinicians to explore participants’ cultural identities, family context, coping strategies, and prior help-seeking experiences to provide culturally responsive care.^32,33,34^ Among South Asian patients specifically, experienced clinicians describe effective care as a process of “two-way education,” in which providers learn about families’ culturally rooted concerns while educating patients about treatment, an approach associated with reduced premature termination of care.^35^ Importantly, these suggestions emerged directly from participants’ lived experiences rather than clinical perspectives, addressing the need for participant-centered research highlighted by Menon et al.^3,31^

Another finding consistently emphasized in the qualitative responses was the need to educate older generations, particularly parents, who continue to shape community attitudes toward mental health. Unlike many previous studies in which intervention targets were proposed by researchers, participants themselves identified parents, elders, faith leaders, and other respected community members as the primary gatekeepers capable of changing community norms. This finding is consistent with previous research demonstrating that older generations strongly influence family and community beliefs regarding mental health.^36^ Older Asian adults are generally less accultured to the United States than younger generations and are more likely to perceive mental health as a sign of weakness.^37^ These generational differences have been described as acculturative family distancing, in which differences in cultural values and communication between immigrant parents and their children increase mental health risk while making emotional concerns more difficult to discuss.^38^ Despite recognition of the need for intervention, programs targeting within South Asian communities remain limited.^39^ A systematic review by Misra et al. found that only 7 of 97 studies on cultural aspects of mental illness stigma evaluated stigma-reduction interventions, and none specifically targeted South Asian parents.^40^ The present findings extend this literature by demonstrating that young South Asian American adults identified older generations, not their peers, as the primary audience for stigma-reduction efforts. Together, these findings suggest that interventions focused solely on young adults are unlikely to produce sustained reductions in stigma and should further engage parents, elders, faith leaders, and other respected community members.

Participants recommended that mental health education extend beyond healthcare settings and be delivered through community institutions, including mosques, temples, churches, and other community organizations. These recommendations align with growing evidence supporting faith-community mental health partnerships. Community-and faith-based interventions have improved mental health literacy, reduced stigma, and increased treatment engagement among minority populations, while faith leaders have been identified as trusted sources of mental health support despite limited formal training.^41–42^ Among South Asian communities, the ROSHNI-2 trial and a recent modified Delphi study further support delivering culturally responsive mental health education through trusted religious and community leaders as an ongoing process rather than a single intervention.^43,44^ Together, these findings support expanding collaborations between healthcare systems, faith leaders, and community organizations to improve culturally responsive mental health education and facilitate access to care among South Asian American communities.

Another important finding was that participants simultaneously emphasized maintaining personal privacy while advocating greater community dialogue surrounding mental health. Rather than representing contradictory views, participants distinguished between protecting the confidentiality of their own care and promoting community-level mental health literacy. This finding extends previous work on selective disclosure, which describes disclosure as context-dependent process rather than a simple choice between openness and secrecy.^45^ Participants explained that privacy involved protecting confidentiality and avoiding community-connected healthcare pathways that might expose treatment, consistent with previous reports describing concerns regarding family reputation, marriage prospects, and breaches of confidentiality.^3,30^ In contrast, openness referred to normalizing mental health discussions through trusted religious and community organizations without requiring personal disclosure.^44^ Together, these findings suggest that stigma reduction efforts should seek to protect individual confidentiality while promoting broader community conversations that normalize mental health and reduce stigma.^45^

Future research should evaluate culturally tailored, community-based interventions targeting parents, elders, and faith leaders while incorporating validated measures of mental health literacy and longitudinal designs to better understand how family attitudes and help-seeking evolve over time. Participant-generated recommendations should also be incorporated into intervention design and evaluation to ensure programs remain culturally relevant and responsive to community needs.

### Strengths and Limitations

This study has several notable strengths, including its mixed-methods design, which integrated quantitative survey data with qualitative open-ended responses. The quantitative survey allowed examination of community-level mental health stigma, barriers to care, and help-seeking attitudes. The qualitative open-ended responses provided deeper insight into participants’ lived experiences, intergenerational differences, and participant-generated recommendations for improving mental health access. The study also included a relatively large sample of South Asian American young adults for both the quantitative portion (*N* = 191) and the qualitative portion (*n* = 144) of the study. The study design also included participants from multiple South Asian ethnic backgrounds, including Indian, Pakistani, Nepali, Bangladeshi, and Sri Lankan communities. The study also focused on South Asian American young adults (ages 18-30) in Texas, a population that remains underrepresented in mental health literature. An additional strength of this study was the inclusion of participant-generated recommendations for improving mental health access, allowing the findings to extend beyond identifying barriers and toward informing culturally relevant intervention strategies.

This study also had several limitations that should be acknowledged. The cross-sectional design precludes causal inference. Additionally, because participants were recruited exclusively from Texas, the findings may not be generalizable to South Asian American communities in other regions of the United States. Because this study was cross-sectional, participants’ residence may have changed following data collection. Therefore, the findings reflect the experiences of South Asian American young adults residing in Texas at the time of study participation and may not represent their current geographic or social contexts. Because study recruitment primarily occurred in larger metropolitan or suburban areas, these findings may not reflect the experiences of South Asian Americans living in rural communities. Furthermore, the recruitment for this research was primarily through university and relatively affluent community networks, so it likely does not reflect South Asian Americans of lower socioeconomic status. While the study sought to include smaller South Asian ethnicities such as Bangladeshi, Sri Lankan, and Nepali South Asian Americans, the sample primarily consisted of Indian and Pakistani participants, which are the ethnicities that primarily make up the South Asian American diaspora in Texas. Women comprised a greater proportion of the sample than men, which may have resulted in greater representation of women’s perspectives and experiences. The surveys were also self-reported, so responses may be subject to recall or social desirability bias. The survey also did not include a questionnaire assessing mental health literacy, limiting the ability to evaluate the relationship between mental health literacy and participants’ attitudes toward mental health and help-seeking. These findings may not be generalizable to older South Asian American adults, whose mental health beliefs, experiences, and mental health literacy may differ substantially. Coding was also conducted by a single researcher, and inter-rater reliability was not assessed. Additionally, the composite measures were developed specifically for this study rather than derived from previously validated scales. Although internal consistency was acceptable, some constructs were conceptually related, and the original family pressure and stigma composites contained overlapping items. A non-overlapping stigma composite was therefore used in multivariable analysis to improve distinction between predictors.

## Conclusion

This mixed-methods study examined mental health stigma and barriers to help-seeking among South Asian American young adults in Texas by integrating quantitative survey findings with qualitative participant perspectives. Although participants reported positive attitudes towards professional mental health care and perceived mental health as important, they continued to experience substantial cultural and social barriers to help-seeking.

In multivariable analysis, mental health stigma and age were significantly associated with help-seeking attitudes after adjustment for family pressure and perceived barriers to care, whereas family pressure and perceived barriers were not independently significant. Qualitative findings further highlighted the influence of family dynamics, intergenerational differences, mental health literacy, privacy concerns, and the need for culturally responsive care, while emphasizing community-based solutions to reduce stigma and improve access.

These findings suggest that improving access to mental health care within South Asian American communities will require interventions beyond the individual. Community-based education targeting parents, elders, faith leaders, and other trusted community members, together with culturally responsive care and provider training in cultural humility, may reduce stigma and improve treatment engagement.

Overall, barriers to help-seeking were shaped less by negative attitudes toward mental health care than by family, community, and cultural contexts. Addressing these broader influences may improve access to care and reduce mental health stigma within South Asian American communities.

## Supporting information

Additional File - English Survey

## Data Availability

The datasets generated and/or analyzed during the current study are available from the corresponding author on reasonable request subject to Institutional Review Board approval.

## Declarations

### Ethics approval and consent to participate

The study was performed in accordance with the ethical principles of the Declaration of Helsinki and was approved by the Sam Houston State University Institutional Review Board (IRB #2026278).

## Consent for publication

Not applicable

## Availability of data and materials

The datasets generated and/or analyzed during the current study are not publicly available because they contain potentially identifiable information from a small and specific study population but are available from the corresponding author on reasonable request, subject to Institutional Review Board approval and applicable data use agreements.

## Authors’ contributions

SK conceived the study with mentorship from XC, designed the study, coordinated participant recruitment, collected the data, performed the quantitative and qualitative analyses, interpreted the findings, and drafted the manuscript. XC supervised the study, provided methodological guidance throughout the project, contributed to study design and interpretation of the findings, critically reviewed and revised the manuscript, and provided overall project oversight. Both authors read and approved the final manuscript.

## Acknowledgements

The authors would like to thank Sam Houston State University College of Osteopathic Medicine Research Committee for selecting this project for funding and for supporting student-led research. We also thank the Medical Summer Scholars Program (MSSP) faculty for their educational programming and support during summer and throughout the research process. We are also grateful to Saima Sheikh for her leadership within the Ahmadiyya Muslim Community, her support with participant recruitment, and to all community members who generously shared their experiences by participating in this study. A special thank you to Rizwan Khan, our friends, family members, classmates, and colleagues who assisted with survey distribution. We further acknowledge Camp Kesem at the University of Texas at Austin and the University of Texas at Austin South Asian community for supporting participant recruitment. Most importantly, we thank all the South Asian American young adults who participated in this study and generously shared their experiences, their willingness to contribute made this research possible.

