## Additional File - English Survey for "Mental Health Stigma and Help-Seeking among South Asian American Young Adults in Texas: A Mixed-Methods Community-Based Needs Assessment"

**Study Title**: Mental Health Stigma and Access to Care Among South Asian American Young Adults

Hello, my name is Sophia Khan, and I am a medical student at Sam Houston State University College of Osteopathic Medicine (SHSU COM). I am conducting a research study under the direction of Dr. Chen to better understand mental health attitudes, stigma, and access to care among South Asian American young adults (ages 18-30) in Texas.

You are invited to participate in this study by completing an anonymous survey. The survey includes questions about mental health attitudes, cultural influences, and barriers to accessing care, as well as basic demographic questions. The survey will take approximately **5-10 minutes** to complete.

To participate, you must be between the ages of 18 and 30, identify as South Asian, and currently reside in Texas.

Your participation in this study is completely voluntary. You may skip any question or stop the survey at any time without penalty.

No identifying information will be collected as part of the survey. At the end of the survey, you may optionally provide your email address if you are interested in participating in a follow-up focus group. This information will be collected separately and will not be linked to your survey responses.

Participants who choose to participate in a follow-up focus group will receive a **$25 gift card** as compensation for their time. Participation in the focus group is completely voluntary.

There are minimal risks associated with this study. Some questions may involve sensitive topics related to mental health. You may skip any questions you are not comfortable answering.

There are no direct benefits to you for participating in this study. However, your responses may help improve understanding of mental health experiences and access to care within South Asian communities.

If you have any questions about this study, you may contact:

Sophia Khan

If you have questions about your rights as a research participant or would like to report a research-related problem, you may contact the SHSU institutional review board at (936) 294-4875 or

Instructions: Please answer the following questions based on your experiences. Participation is voluntary. You may skip any question or stop at any time.

Section 1: Eligibility

1. Are you between the ages of 18-30?

a. Yes

b. No

2. Do you identify as South Asian?

a. Yes

b. No

3. Do you reside in Texas?

a. Yes

b. No

Participants who answer “No” to any eligibility question will not proceed with the survey

Section 2: Demographics

4. What is your age?

a. 18-20

b. 21-23

c. 24-26

d. 27-30

e. Prefer not to answer

5. What is your gender?

a. Female

b. Male

c. Non-binary

d. Prefer to self-describe: ___________

e. Prefer not to answer

6. What is your South Asian ethnicity?

a. Indian

b. Pakistani

c. Bangladeshi

d. Nepali

e. Sri Lankan

f. Other: __________

g. Prefer not to answer

7. What is your current educational/employment status?

a. Undergraduate student

b. Graduate student

c. Employed full-time

d. Employed part-time

e. Other:___________

f. Prefer not to answer

8. What city in Texas do you currently reside in?

a. ___________________

b. Prefer not to answer

Section 3: Mental Health Attitudes

For the following questions, please indicate how strongly you agree or disagree with each statement.

1. Mental health is an important part of overall health

- Strongly Agree
- Agree
- Neutral
- Disagree
- Strongly Disagree

2. I feel comfortable talking about mental health with others

- Strongly Agree
- Agree
- Neutral
- Disagree
- Strongly Disagree

3. Mental health issues are taken seriously by people around me

- Strongly Agree
- Agree
- Neutral
- Disagree
- Strongly Disagree

4. Therapy or counseling is a valid way to address mental health concerns

- Strongly Agree
- Agree
- Neutral
- Disagree
- Strongly Disagree

Section 4: Family and Cultural Influence

For the following questions, please indicate how strongly you agree or disagree with each statement.

5. In my family, mental health is openly discussed

- Strongly Agree
- Agree
- Neutral
- Disagree
- Strongly Disagree

6. My family would support me if I sought mental health treatment

- Strongly Agree
- Agree
- Neutral
- Disagree
- Strongly Disagree

7. There is pressure in my family to maintain a positive image

- Strongly Agree
- Agree
- Neutral
- Disagree
- Strongly Disagree

8. I worry about what others in my community would think if I sought help

- Strongly Agree
- Agree
- Neutral
- Disagree
- Strongly Disagree

Section 5: Stigma

Answer these on a scale of strongly agree to strongly disagree

9. There is stigma around mental health in my community

- Strongly Agree
- Agree
- Neutral
- Disagree
- Strongly Disagree

10. Seeking mental health care is seen as a sign of weakness

- Strongly Agree
- Agree
- Neutral
- Disagree
- Strongly Disagree

11. People in my community are hesitant to talk about mental health

- Strongly Agree
- Agree
- Neutral
- Disagree
- Strongly Disagree

12. Mental health struggles are often kept private within families

- Strongly Agree
- Agree
- Neutral
- Disagree
- Strongly Disagree

Section 6: Help-Seeking Behavior

13. I would feel comfortable seeking professional mental health care

- Strongly Agree
- Agree
- Neutral
- Disagree
- Strongly Disagree

14. I know how to access mental health services

- Strongly Agree
- Agree
- Neutral
- Disagree
- Strongly Disagree

15. I trust mental health professionals

- Strongly Agree
- Agree
- Neutral
- Disagree
- Strongly Disagree

16. I would prefer a provider who understands my cultural background

- Strongly Agree
- Agree
- Neutral
- Disagree
- Strongly Disagree

Section 7: Barriers to Care

Scale: Not a barrier → major barrier

17. Cost or financial concerns

- Not a barrier
- Minor barrier
- Moderate barrier
- Significant barrier
- Major barrier

18. Lack of culturally competent providers

- Not a barrier
- Minor barrier
- Moderate barrier
- Significant barrier
- Major barrier

19. Language barriers

- Not a barrier
- Minor barrier
- Moderate barrier
- Significant barrier
- Major barrier

20. Family expectations

- Not a barrier
- Minor barrier
- Moderate barrier
- Significant barrier
- Major barrier

21. Fear of judgment

- Not a barrier
- Minor barrier
- Moderate barrier
- Significant barrier
- Major barrier

22. Lack of awareness of resources

- Not a barrier
- Minor barrier
- Moderate barrier
- Significant barrier
- Major barrier

23. Religious or cultural beliefs

- Not a barrier
- Minor barrier
- Moderate barrier
- Significant barrier
- Major barrier

Section 8: Open-ended

24. In your opinion, what are the biggest barriers to mental health care in South Asian communities?

__________________________________________________________________

__________________________________________________________________

__________________________________________________________________

__________________________________________________________________

__________________________________________________________________

__________________________________________________________________

__________________________________________________________________

__________________________________________________________________

25. What changes would make it easier for South Asian individuals to seek mental health support?

__________________________________________________________________

__________________________________________________________________

__________________________________________________________________

__________________________________________________________________

__________________________________________________________________

__________________________________________________________________

__________________________________________________________________

__________________________________________________________________

CONTINUE TO NEXT PAGE

Section 9: Optional Focus Group Participation

Contact information provided in this section will be stored separately from survey responses and will not be linked to your survey data during analysis

Participants who complete the follow-up focus group will receive a $25 gift card as compensation for their time

26. Would you be interested in participating in a follow-up focus group?

a. Yes

b. No

27. If yes, please provide your email address below:

______________________________________________________________

Your email will be collected separately and will not be linked to your survey responses.
